# Social Marketing to Enhance Community Empowerment and Ownership for a Successful Implementation of the “Big Catch-Up” in Togo in 2025: A Mixed Method Study

**DOI:** 10.64898/2026.03.11.26348074

**Authors:** Soliou Badarou, Aimé Serge Dali, Kokou Herbert Gounon, Lorraine Shamalla, Amevegbe Kodjo Boko, Xavier Richard Sire, Erinna Corine Dia

## Abstract

**Introduction:** The COVID-19 pandemic disrupted immunization services in Togo, resulting in 69,672 “zero-dose” and 24,846 “under-vaccinated” children by the end of 2023. This study assessed the effectiveness, acceptability, and feasibility of a social marketing approach deployed during the 2025 Big Catch-Up initiative in Togo.

**Methods:** A convergent mixed-methods study was conducted in 17 priority health districts. The quantitative component compared vaccination coverage before and after the intervention using administrative data. Chi-squared test for linear trend compared district-level coverages, and statistical significance was set at p<0.05 for all tests. The qualitative component used in-depth interviews with key informants to collect data, followed by thematic content analysis. The intervention was grounded on the social marketing framework with 4 pillars (4Ps): Product, Price, Place, and Promotion.

**Results:** Coverage increased dramatically: Penta1 from 1% to 64%, Penta3 from 1% to 45%, MR1 from 4% to 50%, and MR2 from 6% to 49% (all p<0.001). Togo ranked 3rd out of 24 African countries for Penta1 progress. The approach demonstrated high community acceptability, with Vaccination Monitoring Committees praised as being culturally appropriate. Key concerns included sustainability and resource constraints.

**Conclusion:** Social marketing proved to be effective for increased community adherence and immunization coverage improvement. However, long-term sustainability requires institutionalization of community structures with domestic funding and continued health system strengthening.

## 1. Introduction

The COVID-19 pandemic has severely disrupted routine immunization services worldwide, creating an unprecedented immunity gap. Between 2019 and 2021, approximately 67 million children missed some or all of their routine vaccinations, marking the largest decline in childhood immunization in three decades [1]. This decline has led to an alarming increase in the number of “zero-dose” children, from 12.9 million in 2019 to 18.1 million in 2021 [2] and 14.3 million in 2024 [3]. In the WHO African Region, vaccination coverage with the third dose of diphtheria-pertussis-tetanus vaccine (DPT3) has fallen from 74% in 2019 to 72% in 2021, jeopardizing decades of progress [4]. This regression has created pockets of vulnerability conducive to outbreaks of measles, polio, diphtheria or yellow fever [5,6].

Sub-Saharan Africa bears the heaviest burden of vaccine-preventable child mortality and accounts for the majority of “zero-dose” children worldwide [7]. Disruptions related to COVID-19 have exacerbated this already precarious situation. In response, WHO, UNICEF, and Gavi, the Vaccine Alliance, launched, in April 2023, the global “Big Catch-Up” (BCU) initiative as an essential immunization recovery plan with three objectives: reach children missed between 2019 and 2022 (partly due to the pandemic) with required antigens, restore vaccination coverage to at least the 2019 levels and strengthen vaccination systems to achieve the Immunization Agenda 2030 goals (IA2030) [8]. Adopted at the World Health Assembly in 2020, IA2030 sets ambitious targets: to achieve 90% vaccination coverage at the national level and 80% in each district by 2030, with a particular focus on reaching “zero-dose” and “under-vaccinated” children [9].

In Togo, the Vaccination Coverage Survey (VCS) conducted in 2024 revealed vaccination coverage rates of 90.3% for DPT3, 80.8% for the first dose of the combined Measles/Rubella vaccine (MR1), and only 57.7% for the second dose of the combined Measles/Rubella vaccine (MR2). Although these rates showed an improvement over previous years, disaggregated analysis revealed significant geographical disparities, with some disadvantaged regions having coverage below 70% for Penta3 [10]. The external review of the EPI conducted in 2019 had already highlighted several structural weaknesses requiring action: shortcomings in the monitoring and tracking of “zero-dose” and “under-vaccinated” children, low community involvement in routine immunization, persistent vaccine hesitancy and rumors, and insufficient knowledge among mothers about the diseases targeted by the EPI, the immunization schedule and the importance of completing the full course of immunization [11]. These shortcomings, exacerbated by disruptions related to the COVID-19 crisis, led to 69,672 “zero-dose” and 24,846 “under-vaccinated” children in Togo, reflecting the global pattern where approximately 67 million children missed some or all of their routine vaccinations during the pandemic[1].

Faced with this critical situation and regional disparities in vaccination coverage, Togo joined global efforts to catch up, restore, and strengthen pre-COVID-19 vaccination coverage levels by implementing the “Big Catch-Up” (BCU) initiative in 2025.

Despite the critical importance of the BCU initiative in addressing the post-COVID-19 immunization gap in Africa, the scientific literature documenting social marketing approaches deployed in this specific context remains extremely limited. This case study on the Togolese experience in this regard has several justifications: It provides evidence on the effectiveness of social marketing approaches in improving immunization demand and services in a West African context. It will help guide future immunization demand programming in Togo and inform national immunization policies [12]. It will also serve as a reference for other African countries facing similar post-COVID-19 vaccination challenges, facilitating the adaptation and replication of approaches that have proven effective [13]. Finally, it will help support resource mobilization efforts to sustain and scale up the approach.

We hypothesized that social marketing can significantly contribute to improving vaccination coverage among “zero-dose” and “under-vaccinated” children. This study aims to evaluate the effectiveness of social marketing approaches in increasing vaccination coverage among “zero-dose” and “under-vaccinated” children in priority districts in Togo in 2025, and assess the implementation process by exploring acceptability and feasibility among various stakeholders.

## 2. Methods

### 2.1. Design and Period

A mixed convergent study combining quantitative and qualitative approaches was used. The quantitative component compared immunization indicators before and after implementation of the accelerated social marketing strategy. The qualitative component used key-informant In-Depth Interviews (IDI) to explore the acceptability and feasibility of the strategy. The study covered two distinct periods: the baseline period (January-June 2025), which is the initial phase of the BCU implementation in Togo, and the intervention period (August-December) that followed the mid-term review and strategic reorientation in July. Data collection occurred in January 2026.

### 2.2. Conceptual Framework

This study was guided by the RE-AIM (Reach, Effectiveness, Adoption, Implementation, Maintenance) framework, a comprehensive model for planning and evaluating public health interventions. Developed by Glasgow et *al*., the RE-AIM framework provides a systematic approach to assess multiple dimensions of intervention success and translation into real-world settings. The framework assessed intervention reach among target populations, effectiveness on vaccination outcomes, adoption by actors, implementation fidelity, and potential for long-term sustainability [14].

### 2.2. Study Setting

The intervention was implemented across four priority health regions of Togo: Grand Lomé, Maritime, Plateaux, and Kara all of which accounted for more than 70% of the country’s zero-dose and under-vaccinated children. Grand Lomé region comprises 2 districts (Agoè-Nyivé and Golfe). Maritime region had 6 priority districts included (Zio, Vo, Yoto, Bas-Mono, Lacs, Avé). **Figure 1**. presents the baseline characteristics of the 4 priority regions selected for intensified intervention. The majority of zero-dose children (n = 36,394) were located in the Golfe district (Figure 1.A), and the majority of under-vaccinated children were located in the Golfe (n = 3,230) and Zio (n = 2,799) districts (Figure 1.B). In total, the intervention targeted 17 districts with 157 health catchment areas identified as the highest priority based on the concentration of zero-dose and under-vaccinated children.

**Figure 1.**
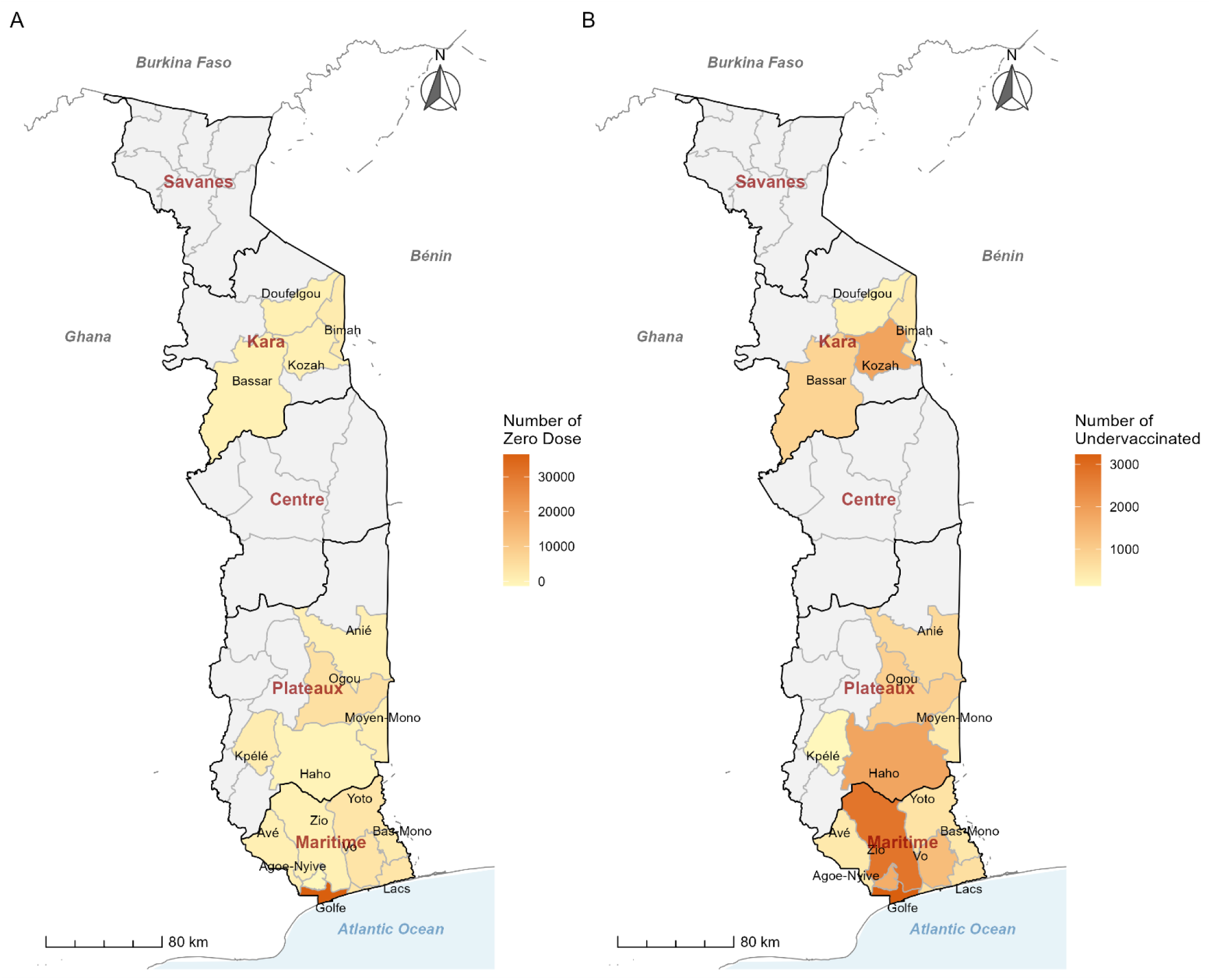
Target regions and districts for “zero-dose” (A) and “under-vaccinated” (B) children for the Big Catch-Up in Togo in 2025.

### 2.3. Intervention: Social Marketing Approach

This approach has demonstrated effectiveness in addressing various public health challenges, including immunization, with evidence showing significant improvements in vaccine coverage when properly implemented [15,16].

In Togo, a comprehensive social marketing strategy was designed and deployed from August to December 2025, following a mid-term review of the BCU that identified key bottlenecks during the initial phase (March-July 2025). This strategy integrated the four pillars of the marketing mix, also known as the 4Ps (Product, Price, Place, and Promotion), to synergistically address service-side, supply-side, and demand-side barriers to immunization (**Figure 2**). This integrated, multi-dimensional approach distinguishes social marketing from isolated health promotion campaigns or service delivery improvements. By simultaneously addressing what is offered (Product), barriers to adoption (Price), accessibility (Place), and community engagement/demand (Promotion), the intervention created mutually reinforcing pathways to behavior change.

**Figure 2.**
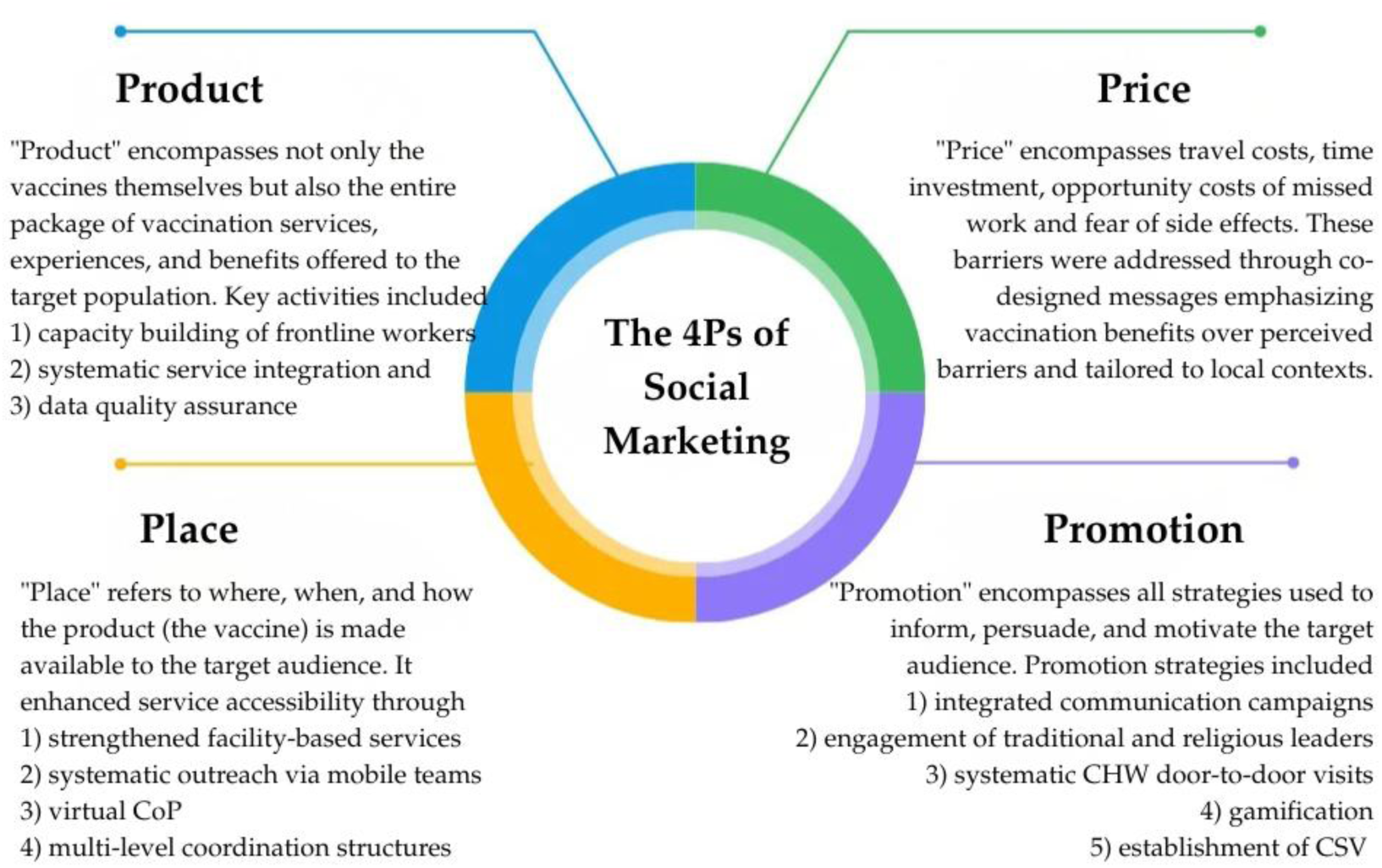
The 4Ps of Social Marketing, adapted from Glasgow et *al* [14]. CoP = Community of Practice, CSV = community-based Vaccination Monitoring Committees.

### 2.4. Quantitative Component

#### Study Population, Sampling Method, and Sample Size

“Zero-dose” children (n=69,672) and “under-vaccinated” children (n=24,846) in priority districts constituted the BCU population. Children aged 0-59 months at the time of the BCU intervention (March-December 2025), residing in Togo and classified as “zero-dose” or “under-vaccinated” based on review of vaccination records or verbal recall if record unavailable. All “zero-dose” and “under-vaccinated” children identified through active case finding during the BCU implementation period were included. The entire target population of 69,672 “zero-dose” and 24,846 “under-vaccinated” children was the denominator for coverage calculation.

#### Data Collection and Analysis

Quantitative data were collected from multiple sources: *DHIS2 (District Health Information Software 2)*; a monthly aggregated vaccination data by antigen, age group, and health facility extracted for the period January-December 2025. DHIS2 is Togo’s national health information system, capturing routine vaccination statistics from all health facilities. *Health facility registers*; vaccination registers at the facility level were reviewed for data completeness and accuracy. *Big Catch-Up specific reporting forms*; custom-designed BCU forms. *Digital dashboard*; real-time data monitoring of vaccination progress, vaccine stock levels, and cold chain functionality. Comparative regional BCU data; aggregated data on BCU performance (Penta1, Penta3 coverage by quarter) for all 23 African countries implementing the initiative, obtained from the Big Catchup Report Dashboard developed by the immunization regional working group (<u>Microsoft Power BI</u>) [17].

Data were analyzed using R version 4.5.0 [18]. Categorical variables were described in terms of frequency and proportion. Vaccination coverage was estimated as a percentage. Chi-squared test for linear trend (Cochran-Armitage test) [19–21] compared district-level coverage across quarters before and during the accelerated intervention period for each antigen (Penta1, Penta3, MR1, MR2). Togo’s quarterly BCU progress was compared with that of other African countries using descriptive statistics and ranking. Statistical significance has been set at p<0.05 for all tests.

#### Operational Definitions

**“***Zero-dose” children* are defined as those who have not received even a single vaccine shot. For operational purposes, GAVI defines zero-dose children as infants who have not received the first dose of diphtheria, tetanus, and pertussis-containing vaccine (DTP1) by the end of their first year of life [22]. “*Under-vaccinated” children* are defined as infants who have not received the third dose of DTP-containing vaccine (DTP3) by the end of their first year of life [22]. *Vaccination coverage* is defined as the proportion of a given population that has been vaccinated in a given time period. It is estimated for each vaccine and, for multi-dose vaccines, for each dose received (e.g., DTP1, DTP2, DTP3). It is usually presented as a percentage [23]. *Priority health areas* are defined as catchment areas of health facilities with the highest burden of “zero-dose” and “under-vaccinated” children within each priority district.

### 2.5. Qualitative Component

#### Inclusion Criteria, Diversification Criteria, and Sampling

Participants were eligible for qualitative interviews if 1) they were directly involved in planning, implementation, supervision, or benefited from the social marketing intervention; 2) and were willing and able to provide informed consent. Purposive sampling was employed and participants were selected to represent geographic and role diversity. Participants were identified through consultation with the country’s Immunization Division and UNICEF’s program officers, who provided lists of key informants at various levels. Based on qualitative research guidelines [24,25] and resource constraints, the target sample size was set to 10 key informants.

#### Data Collection and Analysis

Qualitative data was collected using multiple complementary methods: *In-depth interviews (IDI)*; semi-structured interviews lasting on average 35 minutes were conducted with key informants. Interview guides were developed and covered topics such as the BCU implementation process, the four intervention dimensions, facilitators and barriers encountered, coping mechanisms, perceived effectiveness, acceptability and feasibility, and recommendations. Interviews were conducted in French or local languages, audio-recorded with consent, and supplemented with detailed field notes. *Informal interviews*; with program officers of Unicef and the Ministry of Health. *Document review*: program documents were systematically reviewed, including BCU strategic plans, training materials, communication materials, and supervision reports. This provided contextual information and supported data triangulation. Interviews occurred in private locations convenient for participants (offices, health facilities, and remotely). Participants received no financial compensation but were reimbursed for transport costs when applicable.

Qualitative data were analyzed using thematic content analysis following a systematic process: *Transcription and translation*; all audio recordings were transcribed verbatim within 48 hours of collection. Local language transcripts were translated into French by bilingual translators with public health familiarity and reviewed for accuracy against audio recordings. *Coding*: the analysis was conducted using a hybrid coding approach, combining deductive coding based on the conceptual framework with inductive coding to capture emergent themes from the data. A codebook was developed, and line-by-line coding was performed using RQDA software (version 0.5) [26], with codes systematically applied to relevant text segments. Related codes were grouped into categories and broader themes. *Interpretation:* themes were interpreted in light of the conceptual framework and existing literature. Mechanisms linking intervention components to outcomes were explicated. Findings were reported using thick descriptions with illustrative verbatim quotes.

### 2.6. Ethical and Regulatory Considerations

This case study received ethical approval from the Institutional Review Board of the School of Health Sciences at the University of Lomé in Togo [N° 409/2026/CE-FSS/19/01]. All participants signed a consent form. The confidentiality of the data collected was respected. To ensure anonymity, no information that could identify the participants was used.

## 3. Results

### 3.1. Quantitative Results

#### 3.1.1. Implementation Characteristics

The implementation was assessed through monitoring of key process indicators (**Table 1**).

**Table 1.**
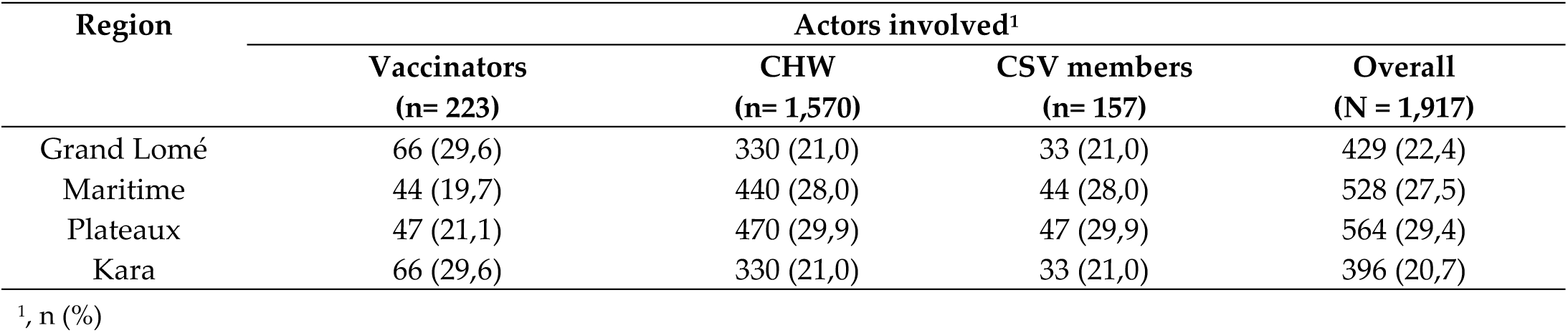
Actors involved in the Big Catch-Up and Social Marketing approach by Category and Region in Togo in 2025.

| Region | Actors involved<sup>1</sup> |  |  |  |
| --- | --- | --- | --- | --- |
|  | Vaccinators<br>(n= 223) | CHW<br>(n= 1,570) | CSV members<br>(n= 157) | Overall<br>(N = 1,917) |
| Grand Lomé | 66 (29,6) | 330 (21,0) | 33 (21,0) | 429 (22,4) |
| Maritime | 44 (19,7) | 440 (28,0) | 44 (28,0) | 528 (27,5) |
| Plateaux | 47 (21,1) | 470 (29,9) | 47 (29,9) | 564 (29,4) |
| Kara | 66 (29,6) | 330 (21,0) | 33 (21,0) | 396 (20,7) |

<sup>1</sup>, n (%)

##### Vaccination Monitoring Committees (CSV) established and functional

The intervention enabled 100% of the target to be achieved. A total of 157 CSV were successfully established across the 157 priority health catchment areas, meeting the objective of one CSV per priority facility. Each CSV comprised 15 members representing diverse community segments, including neighborhood chiefs, CHW, health facility staff, and community leaders. Of the 157 CSV established, 100 % were classified as functional based on criteria including: holding at least 80% of the planned meetings during the intervention period, documented participation in the codesign of vaccination service organization, and active involvement in identifying and referring zero-dose children.

##### Community feedback mechanisms

The CSV served as the primary platform for community feedback and accountability. Across the 157 CSV, a total of 487 community feedback (from suggestions to complaints) were recorded during the intervention period. They were related to vaccination schedule inconveniences (38%), vaccine stockouts or cold chain issues (25%), provider attitudes (18%), long waiting times (12%), and other issues (7%). Of these, 421 (86.4%) were addressed and resolved during subsequent CSV meetings. Unresolved complaints (13.6%) were primarily related to structural issues requiring district or regional-level decisions.

##### Capacity building of vaccinators

A total of 1,917 actors were trained, including 223 vaccinators (11,6%), and 1,570 CHW and community relays (81,9%). All participants received training in at least two evidence-based communication techniques. Religious leaders additionally received a briefing on the intervention principles. **Table 1** presents the distribution of trained actors by category.

#### 3.1.2. Evolution of Vaccination Coverage

##### Temporal Distribution of Vaccination Coverage

The accelerated social marketing intervention resulted in dramatic improvements in vaccination coverage for all tracer antigens. **Figure 3.A** presents the evolution of Penta1 and Penta3, and **Figure 3.B** presents the evolution of MR1 and MR2 coverage from January to December 2025.

**Figure 3.**
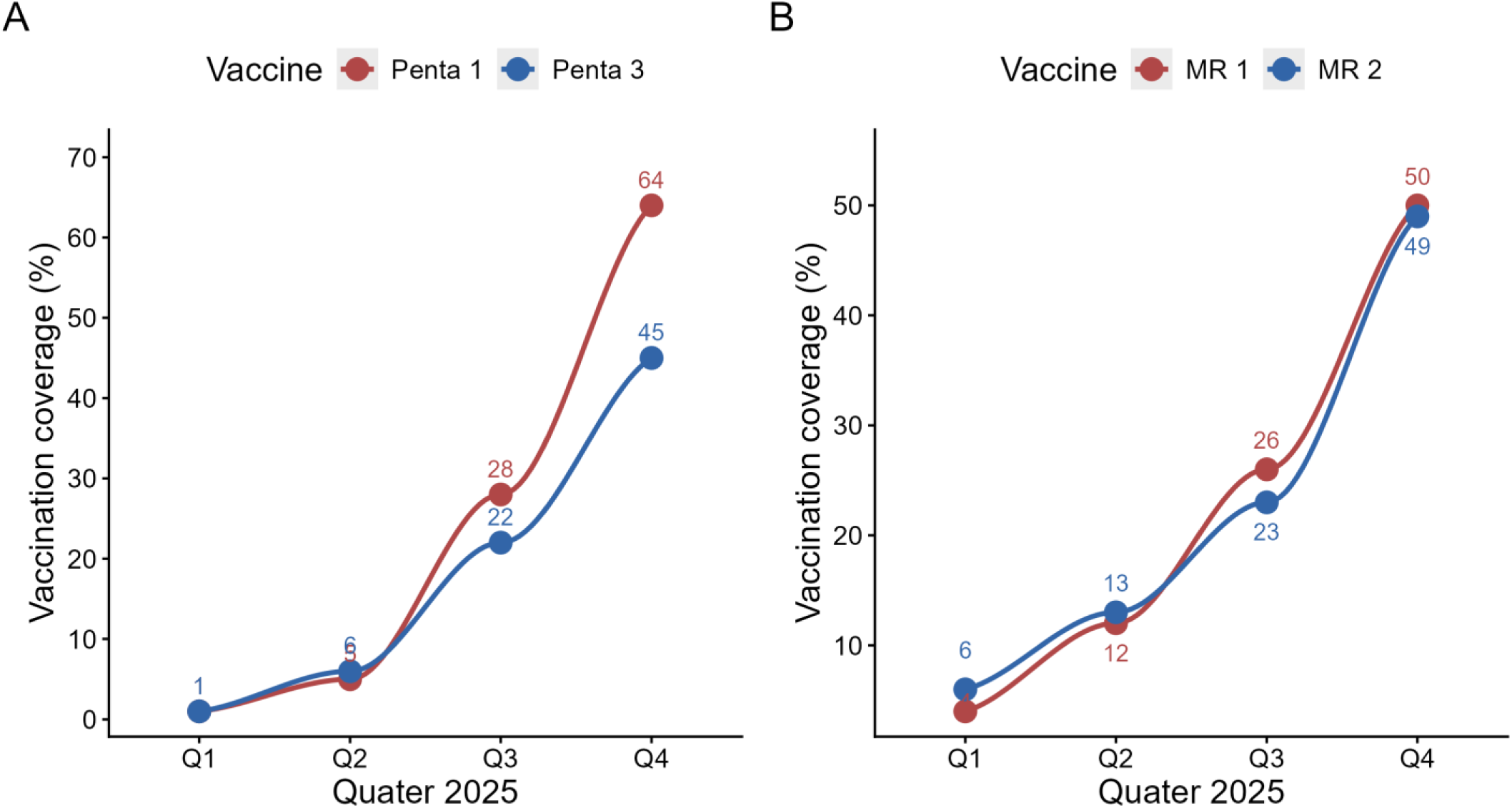
Temporal distribution of Vaccination Coverage of Penta (A) and MR (B) in Togo in 2025.

Penta1 coverage increased progressively from 1% in Q1 (pre-acceleration phase) to 64% by Q4. The August-December phase alone contributed 59 percentage points of the 63% annual progress, representing 94% of the total annual gain achieved in just 5 months. The analysis revealed a statistically significant inflection in the trend (p<0.001). For Penta3 coverage, a similar acceleration pattern was observed. Coverage rose from 1% (Q1) to 45% (Q4). The acceleration phase contributed 40 of 44 percentage points, representing 91% of the total annual gain. The inflection in the trend was statistically significant (p<0.001).

MR1 coverage also showed dramatic gains, increasing from 4% (Q1) to 50% (Q4). The August-December period accounted for 38 of 46 percentage points gained, representing 83% of the total annual gain. Pre and post-intervention increase was statistically significant (p<0.001). MR2 demonstrated substantial improvement from 6% (Q1) to 49% (Q4). The intervention period contributed 36 of the 43 percentage points gained, representing 84% of the total annual gain. The increase in the trend was statistically significant (p<0.001).

##### Spatial Distribution of Vaccination Coverage by Health District

**Figure 4** shows progress in vaccination coverage in Penta1 and **Figure 5**, in Penta3 by district and by quarter. For Penta1, there was a clear increase in vaccination coverage between Q2 (Figure 4.B) and Q3 (Figure 4.C), with the best performance in the health districts of Zio, Bassar and Doufelgou (100%) in Q4 (Figure 4.D). For Penta3, there was also a clear increase in vaccination coverage between Q2 (Figure 5.B) and Q3 (Figure 5.C), with the best performance in the health districts of Agoè-Nyive (100%), Doufelgou (94%) and Avé (82%) (Figure 5.D).

**Figure 4.**
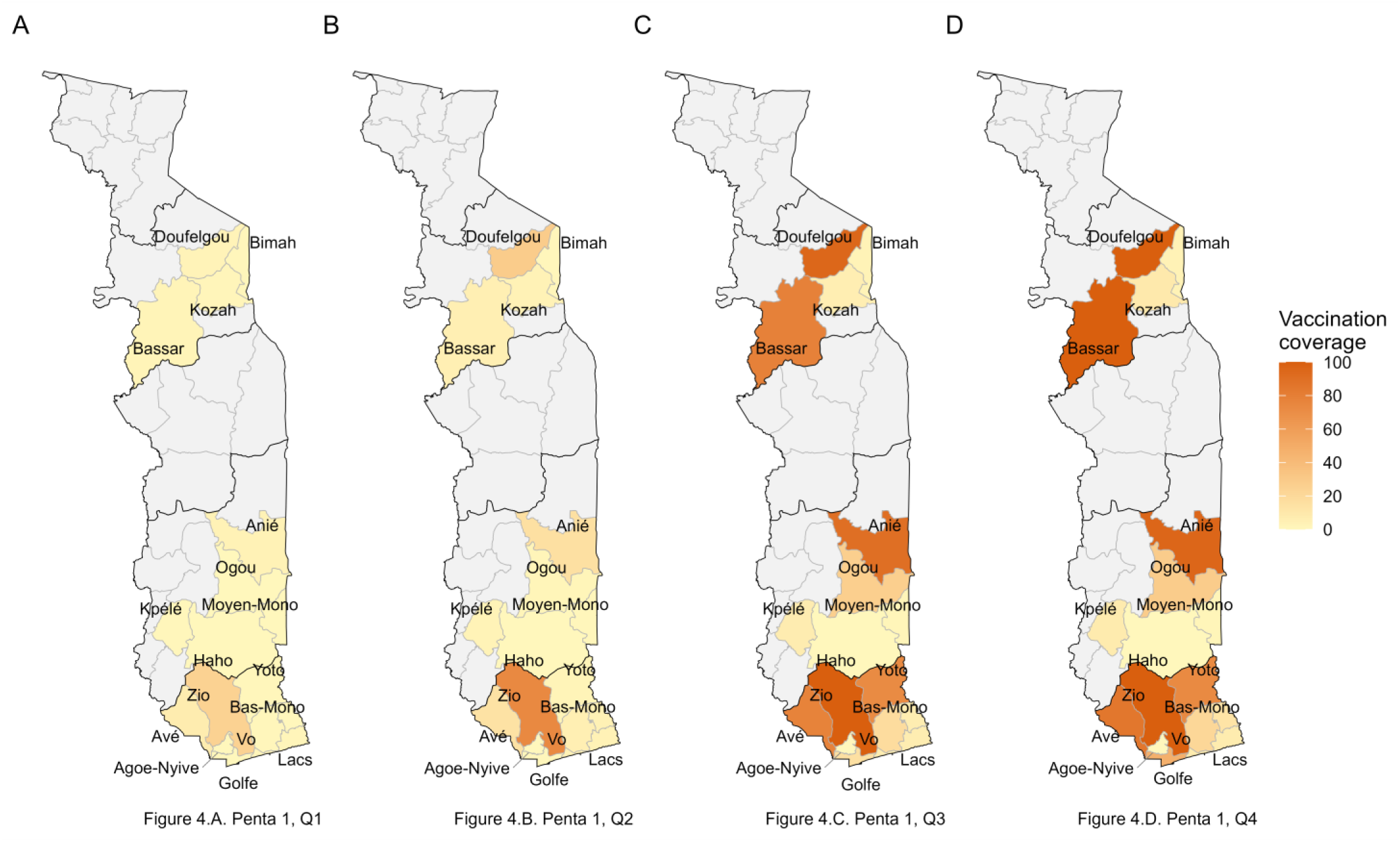
Spatial distribution of Vaccination Coverage of Penta 1 by quarter in Togo in 2025.

**Figure 5.**
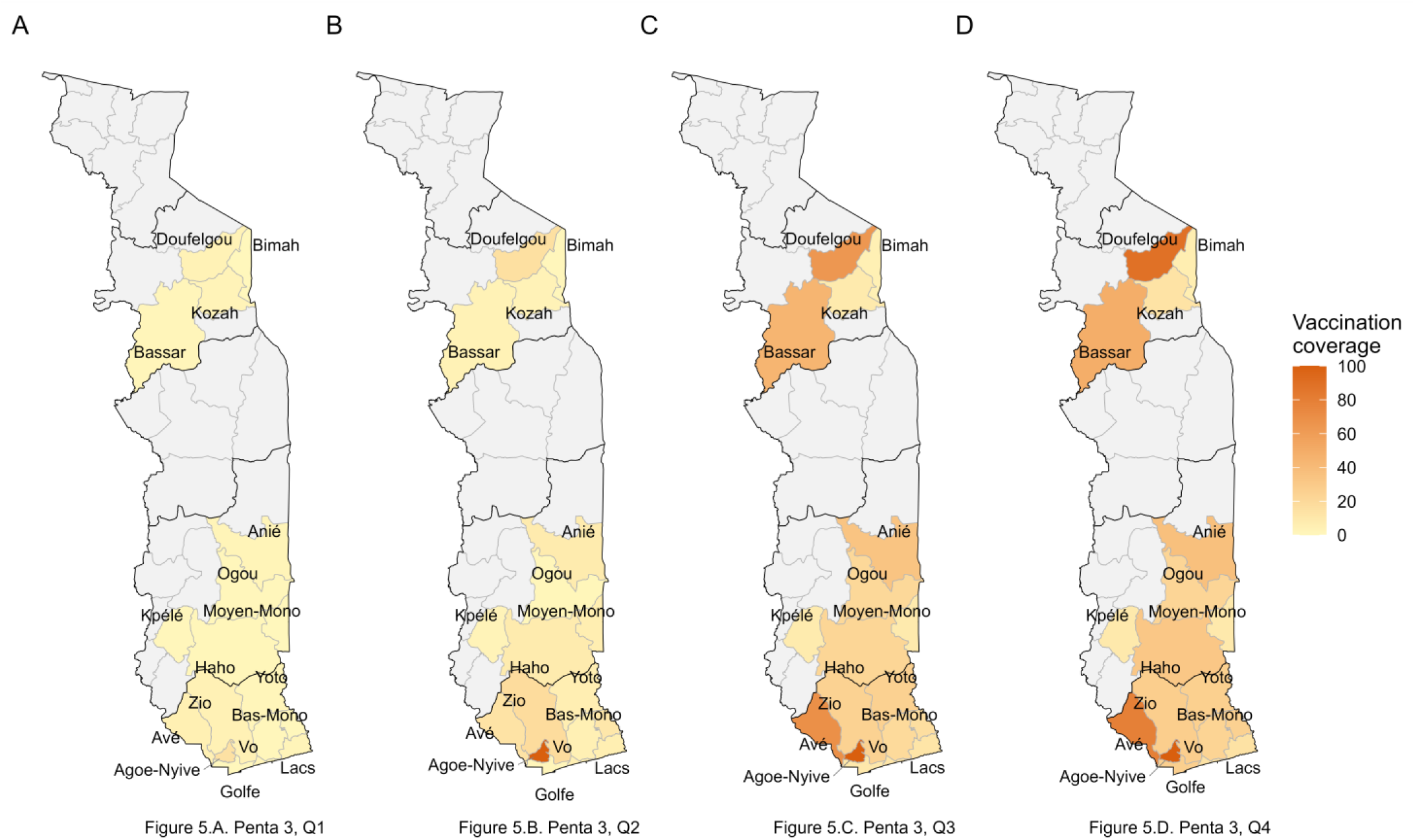
Spatial distribution of Vaccination Coverage of Penta 3 by quarter in Togo in 2025.

#### 3.1.3. Comparison of Togo’s Performance with Other African Countries

Togo’s performance in the BCU initiative was benchmarked against 23 other African countries implementing the program. **Figure 6** presents the comparative regional performance for Penta 1 and Penta 3 coverage progression.

**Figure 6.**
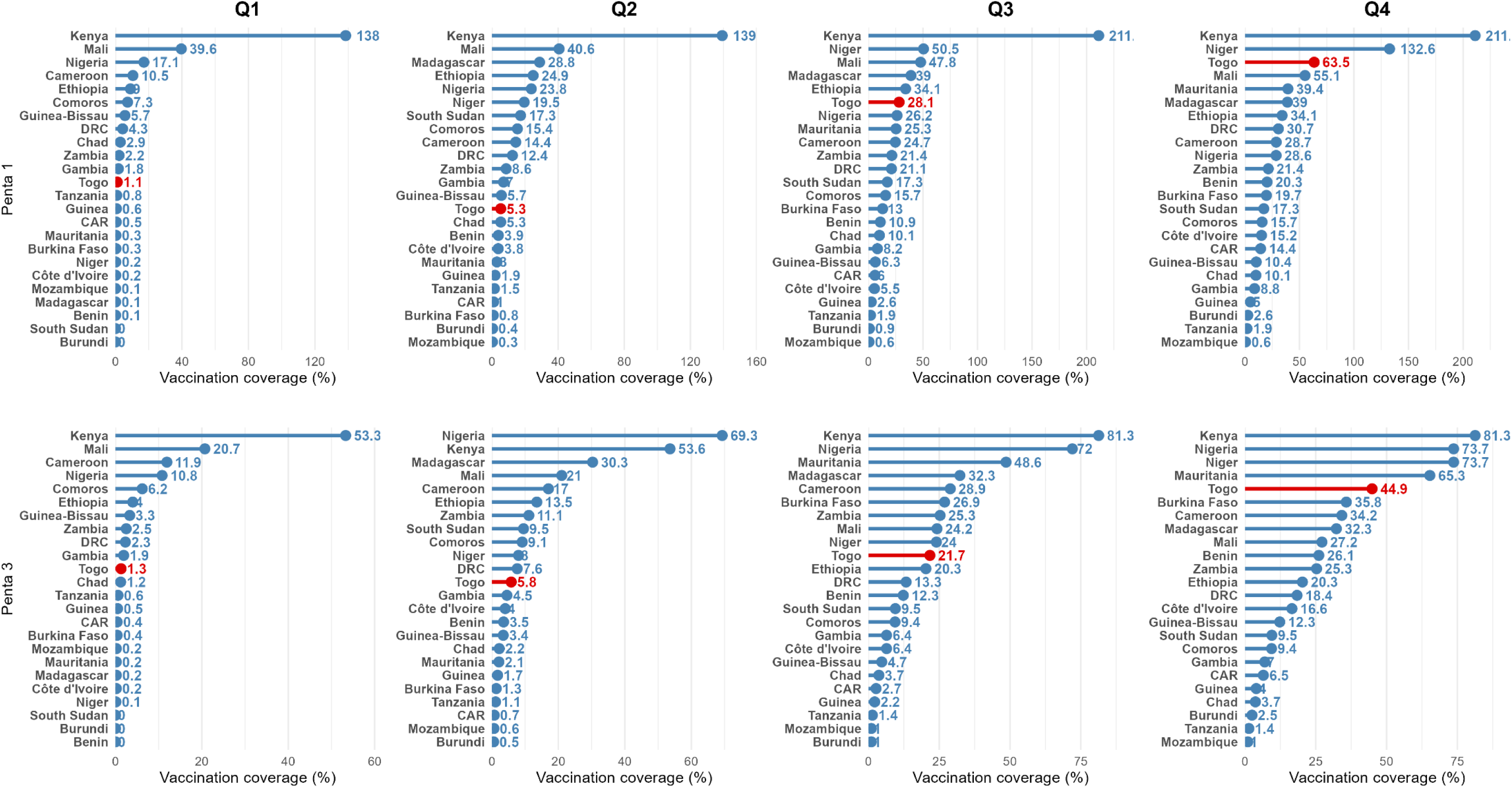
Comparative Performance of African Big Catch-Up Countries for Penta 1 and Penta 3 Coverage in 2025.

For the Penta 1 progression (January-December 2025), Togo ranked 3rd of 23 African BCU countries, with 63.5% coverage in December 2025. Notably, Togo dramatically improved from 12th place in Q1 to 3rd place in Q4. Togo’s half-yearly gain of 58.2 percentage points between Q2 and Q4 was the second-highest acceleration among all countries, exceeded only by Niger (113.1 points).

For Penta 3 progression (January-December 2025), Togo ranked 5th with 44.9% coverage, placing in the middle tier of performers. Kenya led dramatically with 81.3%, followed by Niger and Nigeria (both 73.7%). Togo’s Penta3 ranking lagged behind its Penta 1 ranking (number 5 vs number 3). However, strong acceleration was observed during Q2-Q4 with a jump from 5.8% (Q2) to 44.9% (Q4), placing the country in the 5th-highest among accelerating countries.

### 3.2. Qualitative Results

#### 3.2.1. Sociodemographic Characteristics of Respondents

Of 10 individual interview participants, 2 was females and 8 males. Age ranged from 35 to 54 years; the median age was 44 years (IIQ = 40.75 −49.75) and the median years of experience as health professional was 11 years (IIQ = 7 – 16). Professional roles included members of CSV (n=2), health promotion focal point (HPFP) (n=2), health facility heads (HFH) (n=1), immunization focal point (IFP) (n=2), prefectural health director (PHD) (n=1), member of COGES (Management Committees) (n=1) and Beneficiary (n=1). Geographic diversity was achieved with participants from 5 health districts: Agoè-Nyivé, Bas mono, Golfe, Vo and Yotto.

**Table S1** provides an extensive overview of recurring themes and examples of quotes regarding the intervention’s acceptability, feasibility, barriers, and facilitators.

#### 3.2.2. Acceptability

The intervention demonstrated high levels of community acceptability across regions, with stakeholders at all levels expressing positive perceptions of the approach. The most recurring themes were: CSV as a culturally appropriate mechanism, Engagement of religious/traditional leaders, Gamification and recognition, and Behavioral change.

##### CSV as culturally appropriate engagement mechanism

The establishment of CSV was praised as a culturally appropriate mechanism for community engagement that respected local decision-making structures while introducing systematic accountability. A health promotion focal point in Bas mono explained:

*“The community appreciated the CSV because they feel that their loved ones and parents are raising their awareness about something they expected to have side effects from. They see that it is the community that is reaching out to them to tell them that it is good to get vaccinated.”* (IDI04-IFP-Golfe).

##### Religious and community leader engagement appreciated

The involvement of religious and community leaders was widely accepted and effective. A health promotion focal point in Bas mono district shared:

*“Some pastors asked children to bring their vaccination records to church. They checked them and found that a certain number of children in their communities had not been vaccinated. They see that there is an CHW in the area and entrust the children to them*.

*As for the Vodou priests, it is in the convents and gatherings for ceremonies*.

*For imams, it is at the mosques. In each area, the imams were invited. For example, in Attitongon, they passed on the information to the mosque. Some imams brought children to the health facilities. They found, convinced (…) and accompanied the children.”* (IDI02-HPFP-Bas-mono).

##### Gamification and recognition ceremonies acceptable

The performance-based recognition system and public ceremonies generated mixed reactions. High-performing districts and facilities appreciated the visibility and motivation. A health promotion focal point stated:

*“Because we won a trophy with a medal. We came to present it at the district level*.

*At first, all the leaders were happy. Everyone on the CSV committees was happy with the activities they had carried out. When they saw the trophy, the village chiefs were very happy.”* (IDI02-HPFP-Bas-mono).

However, low-performing districts expressed some concern about public discomfort. But overall, acceptability was high when: recognition emphasized learning rather than only competition. A Prefectural Health Director confided:

*“When it comes to social marketing, sharing results should not be seen as something that will upset anyone. No, we are all here together, sharing our experiences. It is a place where those who are less successful can learn from the experiences of those who have done good work in the field to improve coverage.”* (IDI05-PHD-Vo)

##### Behavioral change

Many behavioral changes were reported during and after the activities, with special mention of adherence to the vaccination schedule and a decrease vaccine hesitancy. A member of CSV stated:

*“Hesitancy and other issues have changed. This has allowed us to have a larger number. Above all, adherence to schedules has also been important.”* (IDI01-CSV-Agoè-nyivé)

#### 3.2.3. Operational Feasibility

The intervention was generally perceived as operationally feasible, though implementation challenges required adaptive solutions. The most recurring themes were: Simultaneous approach of awareness raising, research, and vaccination; Cascade training and Home visits.

##### Simultaneous approach of awareness raising, research, and vaccination

In this context, it became apparent that awareness-raising efforts were not limited to a single task but were part of a continuum of integrated activities aimed at maximizing the impact of vaccination campaigns within the communities visited. A member of the COGES explained:

*“During vaccination, if we arrive in a region, we vaccinate them there, in the community. We set up a place under the trees or at the notable’s or chief’s house, and vaccinate them there. For some, we direct them to the Peripheral Health Unit. But there are certain areas that are a little far from the Peripheral Health Unit, so whenever we raise awareness, we vaccinate them there.”* (IDI06-COGES-Yoto)

##### Training implementation feasible with cascading approach

Training 1,917 actors within the compressed timeframe was achieved through a cascading model: Master trainers trained focal points, who in turn trained district teams, who eventually trained facility-level actors and CHWs. A health facility head described:

*“… we were also trained at the beginning. There were ASC training sessions that followed. (…) we also trained them before sending them out into the field. For the community, we had an initial training session for all community members. (…) some members were trained as CSVs.”* (IDI03-HFH-Golfe).

##### Home visits operationally demanding but desirable

Conducting such home visits over the project implementation period required significant CHW effort. This logistical and human aspect of home visits has been described as particularly demanding, but also perceived as essential. A beneficiary reported:

*“For my part, I was informed by a relative who is a community health worker. (…) He came to our house with two of his colleagues, who asked us if our children had been vaccinated. They explained the benefits of vaccination to us.”* (IDI07-Beneficiary-Yoto).

#### 3.2.4. Perceived Barriers to Implementation

Despite overall success, several barriers hindered optimal implementation. The most recurring themes were: Geographical and infrastructural barriers, Organizational and resource barriers, Community and cultural barriers, and Sustainability concerns

##### Sustainability concerns

Although some CSV members continue to do their work despite the end of the project, participants nevertheless raised substantial concerns about the sustainability of activities, pointing out that, despite the positive effects observed in the short term, CSV may not be sustainable without the continued commitment of local authorities. This has led to active dialogue with mayors to explore mechanisms for long-term institutional and financial support. A Prefectural Health Director stated:

*“At present, we have all seen the impact on our activities. So, as of today, what we have done most is to approach local authorities, especially mayors, to see how they can help us to keep our activities going. (…) We are in discussions, and the grievances are on the mayor’s table.”* (IDI05-PHD-Vo)

##### Geographical and infrastructural barriers

Hard-to-reach areas, displaced people faced significant access challenges as well as rainy weather conditions. Geographical barriers were consistently mentioned by actors as major logistical challenges. A Prefectural Health Director added:

*“The main difficulty was that the activity was carried out in rainy weather. And there were difficulties, especially in reaching certain areas.”* (IDI05-PHD-Vo)

##### Organizational and resource barriers

Some participants expressed organizational and human resource concerns, particularly regarding the insufficient number of community workers available to carry out a sufficient number of visits, which could limit the number of children actually vaccinated in certain areas. A health promotion focal point worried:

*“But we also noticed that there weren’t enough CHWs assigned. (…) There were almost 30 CHWs with community relays. Only 15 were selected. So that bothered us a little. But we tried to manage it that way*.

*And now we can see the number of vaccination team visits. (…) Because in some districts, this has somewhat hindered the number of children vaccinated. Because we have given six visits. And if the CHWs stick to these six visits, there will certainly be children who remain unvaccinated. We need to increase (…) the number of visits by vaccinators.”* (IDI02-HPFP-Bas-mono).

#### 3.2.5. Perceived Facilitators

Multiple factors are perceived to have facilitated successful implementation:

##### Awareness raising

Awareness-raising activities helped to change beneficiaries’ perceptions of vaccination, not only by providing clear information about its benefits, but also by tailoring the message to families’ living conditions and reducing individual apprehensions through personalized discussions. A beneficiary noted:

*“As my two children were not vaccinated, they offered us the vaccination. (…). They still managed to convince me of the benefits of vaccination and how important it is for our children’s lives. (…) When it comes to the message about vaccination, when you go to the hospital, the message gets across. However, when you are at home, and people come to visit, they explain things to you in a different way*.

*And even when you are afraid, the fear dissipates.”* (IDI07-Beneficiary-Yoto)

##### Codesign approach and community engagement

Unlike top-down campaigns, the codesign methodology genuinely engaged communities in solution design, generating ownership. A health facility head contrasted:

*“(…) We held meetings with the community. They were the ones who came up with key ideas for mobilizing the population around vaccination. (…) It is an advantage to involve people who live in the community (…) they understand that their health is in their own hands.”* (IDI03-HFH-Golfe).

##### Strong commitment from Government

Close collaboration between the UNICEF technical team and the government leadership at the operational level created synergy. An Immunization focal point noted:

*“UNICEF, which spearheaded the social marketing initiative, collaborated with the government. (…) The collaboration consisted of (…) pooling the various services (…) by inviting other sectors that UNICEF considered influential for the success of social marketing, notably the health promotion division and the immunization division of Ministry of health (…). This was crucial.”* (IDI04-IFP-Golfe).

### 3.3. Integration of Quantitative and Qualitative Results

Triangulation of quantitative performance data with qualitative actor experiences revealed convergent and complementary findings.

#### Convergence on community engagement effectiveness

Quantitative data showed 100% of CSV were functional, and 86.4% of community feedback was addressed. Qualitative data explained *why* these mechanisms worked (cultural appropriateness of co-creation, respect for local decision-making) and *how* they translated into outcomes (generating ownership, enabling context-specific solutions, creating accountability).

#### Complementarity on capacity building impact

Quantitative results documented 1,917 actors trained. Qualitative findings added depth, revealing that training such number of actors required a cascade training strategy. Furthermore, qualitative data revealed that training’s impact extended beyond individual skill acquisition to creating a community where vaccinators, CHWs, and leaders mutually reinforced vaccination messages.

#### Sustainability insights from divergent perspectives

Quantitative data showed high CSV functionality during intervention (100%). Qualitative data revealed sustainability concerns; anticipated decline when external support ends and need for continued modest financial support.

The integrated mixed-methods design thus provided a comprehensive, nuanced understanding unattainable through either method alone: quantitative evidence of effectiveness, qualitative insights into mechanisms, acceptability, feasibility, contextual factors, and sustainability considerations.

## 4. Discussion

The quantitative results revealed substantial progress: Penta1 vaccination coverage rose from 1% in Q1 to 63.5% in Q4, and Penta3 coverage rose from 1% in Q1 to 44.9% in Q4, with a marked acceleration during the intensive intervention phase. Similarly, MR1 progressed from 4% to 50%, and MR2 from 6% to 49%. Regionally, Togo ranked 3rd out of 24 African countries for Penta1 progress and 5th for Penta3. Qualitative results show strong community acceptance of the approach, particularly thanks to the CSV, which served as a culturally appropriate mechanism for community engagement. The involvement of religious and traditional leaders, combined with home visits and the simultaneous awareness-raising, research, and vaccination approach, has led to notable behavioral changes, including reduced vaccine hesitancy and better adherence to the vaccination schedule.

We reported coverage increasing from 1% to 64% for Penta 1 and from 1% to 45% for Penta 3 in six months. Similar progress was reported in Angola, where an approach combining community mobilization and the involvement of traditional leaders increased DPT1 coverage from 75.8% to 90.2% and DPT3 coverage from 67% to 84% in one year [27]. This similarity in the progression of vaccination coverage suggests that intensified social mobilization strategies produce comparable effects in different African contexts. Rapid progress during the acceleration phase was also reported in Ethiopia, where the relaunch of the BCU in February 2025 resulted in the vaccination of nearly 100,000 children in just two months, increasing coverage from 16% to 24% of the target population [28]. These rapid accelerations demonstrate that well-coordinated strategic intensification can produce substantial results in a relatively short period of time.

We reported high acceptability of CSV as a mechanism for community engagement. Similar results have been reported in Nigeria, where the Community Health Influencers, Promoters, and Services (CHIPS) program adopted a similar approach to community mobilization with convincing results: more than 4 million eligible children vaccinated between March 2021 and January 2023, including more than 700,000 who received Penta3 [29]. This could be explained by community ownership of health interventions, a fundamental principle for reaching marginalized populations.

The involvement of religious and traditional leaders has proven crucial to our approach. A systematic review of immunization demand strategies in Africa reported similar results, showing that community mobilization using indigenous communication channels (town criers, religious leaders) significantly increases vaccination coverage [30]. This could be explained by the influence of these leaders as reliable sources of information, which helps to counter misinformation and build trust in vaccination services, particularly in rural areas where their authority is predominant.

The simultaneous approach of awareness-raising, screening and vaccination was an important operational innovation that maximized efficiency. Carcelen et *al* reported, in a scoping review, a similar optimization approaches, notably the PIRI (Periodic Intensification of Routine Immunization) approach in India, which was organized for seven consecutive days each month [31]. By avoiding the delay between identification and vaccination, these approaches reduce dropouts and improve completion rates.

The use of the 4Ps of social marketing (Product, Price, Place, Promotion) in our vaccination context generated substantial increases of 1% to 45% for Penta3. Zheng et *al* in China in 2023 used the same social marketing strategy for COVID-19 vaccination and reported a significant reduction in vaccine hesitancy from 52% to 3.1% and an increase in vaccination coverage to 94.8% [15]. Similarly, Hong et *al* in South Korea in 2023 used the 4Ps strategy of social marketing and reported 94.8% coverage for the second dose and 71.3% for the third dose of COVID-19 vaccination [32]. These international results validate the applicability of this strategy to different contexts and populations.

The programmatic implications are manifold. For Togo, it is essential to capitalize on established CSVs and institutionalize them in the routine health system with sustainable government funding. The codesign and community engagement approaches should be extended to other health interventions to maximize investment in these structures. Strengthening the health information system, particularly the quality of immunization data, must continue to ensure accurate monitoring and data-driven planning. For other African countries facing similar challenges, this Togolese experience offers transferable lessons.

### Strengths and Limitations

This evaluation has several methodological and conceptual strengths. The use of a mixed design combining quantitative and qualitative analyses provided a holistic understanding of the intervention. This methodological triangulation strengthens the validity of the conclusions. The evaluation was based on the integrated RE-AIM conceptual framework (Reach, Effectiveness, Adoption, Implementation, Maintenance). This structured theoretical approach enabled us to evaluate not only the effectiveness of the intervention, but also its reach, adoption by providers, fidelity of implementation, and potential for sustainability. This multidimensional evaluation goes beyond simple measures of vaccination coverage. The inclusion of 17 priority health districts in four regions representing more than 70% of the country’s “zero-dose” and “under vaccinated” children ensures substantial geographical representativeness. This targeted approach, based on the burden of disease, optimizes resource allocation while generating data that can be generalized to areas with a high prevalence of unvaccinated children. The comparative analysis with 23 other African countries implementing the BCU provides an essential regional context for assessing Togo’s performance. This regional perspective allows Togo’s progress to be situated on a performance gradient and guides cross-country learning for continuous improvement.

However, certain methodological limitations must be considered. The absence of a control group limits the strict causal attribution to our intervention. The design adopted cannot completely exclude the influence of confounding factors such as competing interventions that may have contributed to the improvements observed. The short follow-up period does not allow for an assessment of the long-term sustainability of coverage gains or the sustainability of CSV. The quality of administrative data is a significant limitation. Despite efforts to triangulate and improve data quality, inconsistencies or underreporting may persist in some health facilities.

## 5. Conclusions

The social marketing approach deployed in Togo as part of the Big Catch-Up has proven effective in rapidly reaching zero-dose and under-vaccinated children, thereby helping to reduce the post-pandemic immunization gap. The success of this intervention is based on community engagement, the commitment of local leaders, strengthened multilevel coordination, and the synergistic integration of the four pillars of social marketing. For these gains to translate into sustainable progress towards the goals of the Immunization Agenda 2030, sustained political and financial commitment from the government, institutionalization of community mechanisms, and continuous strengthening of primary health systems are imperative. The Togolese experience confirms that vaccine equity in Africa is achievable, but requires political will, substantial domestic investment, and participatory approaches that place communities at the heart of vaccination strategies.

## Supplementary Materials

Supplementary information are available in Table S1.

## Author Contributions

Conceptualization, SB and ASD; methodology, KHG and ASD; software, KHG; validation, SB, XRS and ECD; formal analysis, KHG; resources, SB, XRS and ECD; data curation, KHG and ASD; writing—original draft preparation, KHG; writing—review and editing, KHG, LS, ASD, SB, XRS and ECD; supervision, SB; project administration, ASD; funding acquisition, SB, XRS and ECD. All authors have read and agreed to the published version of the manuscript.”

## Funding

This research was funded by Unicef, under the grant number SC220798, GAVI. The APC was not funded.

## Institutional Review Board Statement

The study was conducted in accordance with the Declaration of Helsinki, and approved by the Institutional Review Board of the School of Health Sciences at the University of Lomé in Togo (N° 409/2026/CE-FSS/19/01).

## Informed Consent Statement

Informed consent was obtained from all subjects involved in the study.

## Data Availability Statement

One part of the original data presented in the study is openly available in the Big Catchup Report Dashboard developed by the immunization regional working group (<u>Microsoft Power BI</u>) at https://app.powerbi.com/view?r=eyJrIjoiMWE2M2M3M2UtZDF-hOC00MzE5LThmNjQtND-QwNmY4ZGY2NGEzIiwidCI6ImY2MTBjMGI3LWJkMjQtNGIzOS04MTBiLT-NkYzI4MGFmYjU5MCIsImMiOjh9. The other part of data presented in this study are available on request from the corresponding author due to restricted access to Ministry of Health staff only.

## Acknowledgments

The authors would like to thank all participants in this study. During the preparation of this manuscript, the authors used ChatGPT as a writing assistance tool and DeepL as a rephrasing tool. The authors have reviewed and edited the output and take full responsibility for the content of this publication.

## Conflicts of Interest

The authors declare no conflicts of interest. The funders had no role in the design of the study; in the collection, analyses, or interpretation of data; in the writing of the manuscript; or in the decision to publish the results.

## Abbreviations

The following abbreviations are used in this manuscript:

BCU: “Big Catch-Up”
CAR: Central African Republic
CHW: Community Health Worker
CoP: Community of Practice
COGES: Management Committee
CSV: French acronym for Vaccination Monitoring Committee
DHIS2: District Health Information Software 2
DTP1/DTP3: Diphtheria-Tetanus-Pertussis 1/3 (first/third dose)
DRC: Democratic Republic of the Congo
VCS: Vaccination Coverage Survey
EPI: Expanded Program on Immunization
GAVI: Global Alliance for Vaccines and Immunization, The Vaccine Alliance
HFH: Health Facility Head
HPFP: Health Promotion Focal Point
IA2030: Immunization Agenda 2030
IDI: In-Depth Interviews
IFP: Immunization Focal Point
MR1/MR2: Measles-Rubella 1/2 (first/second dose)
Penta1/Penta3: Pentavalent vaccine 1/3 (first/third dose)
PHD: Prefectural Health Director
PIRI: Periodic Intensification of Routine Immunization
RE-AIM: Reach, Effectiveness, Adoption, Implementation, Maintenance

## Notes

### Competing Interest Statement

The authors have declared no competing interest.

### Author Declarations

Institutional Review Board of the School of Health Sciences at the University of Lome in Togo gave ethical approval for this work [Number 409/2026/CE-FSS/19/01].

