## Supplementary Table S1 for "Social Marketing to Enhance Community Empowerment and Ownership for a Successful Implementation of the “Big Catch-Up” in Togo in 2025: A Mixed Method Study"

**Table S1.** Selection of verbatim quotes illustrating the identified themes

| Themes | Quotes |
|---|---|
| **Acceptability** | |
| Engagement of religious/traditional leaders | Some pastors asked children to bring their vaccination records to church. They checked them and found that a certain number of children in their communities had not been vaccinated. They see that there is an CHW in the area and entrust the children to them. As for the Vodou priests, it is in the convents and gatherings for ceremonies. (**IDI02-HPFP-Bas-mono**)<br><br>For imams, it is at the mosques. In each area, the imams were invited. For example, in Attitongon, they passed on the information to the mosque. Some imams brought children to the health facilities. They found, convinced (...) and accompanied the children.<br>Once these leaders have been reached, all their followers are reached, and this has enabled us to get these religious leaders to call us to say that there are two or three children with them and the health team will vaccinate these children.<br>On Sundays, we go to church, the pastor or priest gives us some time and we explain to the community. We explain social marketing and the benefits of vaccination. Some people benefit immediately from vaccination, as we set up a vaccination team in front of the church. (…) We used the same principle and tried it out in mosques, churches and even with traditional priests. (**IDI05-PHD-Vo**)<br><br>There is an association of charlatans. The representative who is among us went to talk to the other charlatans. He also spoke with the adepts. The pastors also have their association here. This helped us to enter the churches to search for those who had been left behind. (**IDI06-COGES-Yoto**) |
| CSV as a culturally appropriate mechanism | The community appreciated the CSV because they feel that their loved ones and parents are raising their awareness about something they expected to have side effects. They see that it is the community that is reaching out to them to tell them that it is good to get vaccinated. (**IDI04-IFP-Golfe**)<br><br>We still believe that the involvement of traditional chiefs and religious leaders from the outset of social marketing really made a difference (...) there was active participation, which facilitated acceptance of the project and dissemination of the messages.<br>This proximity really helped to build trust within the community. Trust in the health system began to emerge, which was a good thing. (**IDI05-PHD-Vo**)<br><br>The fact that he is a close friend, someone we know well, makes it easier for us to relate and talk to each other. (...) So if my children are ill, I find it easier to refer to him because we are close. (**IDI07-Beneficiary-Yoto**) |
| Gamification and recognition | When it comes to social marketing, sharing results should not be seen as something that will upset anyone. No, we are all here together, sharing our experiences. It is a place where those who are less successful can learn from the experiences of those who have done good work in the field to improve coverage. (**IDI05-PHD-Vo**)<br><br>They too (...) did their best anyway. It depends on the area. And it also depends on the resources available. We can encourage them and tell them to do better. (**IDI06-COGES-Yoto**)<br><br>Because we won a trophy with a medal. We came to present it at the district level.<br>At first, all the leaders were happy. Everyone on the CSV committees was happy with the activities they had carried out. When they saw the trophy, the village chiefs were very happy. Next time won't be easy. (**IDI02-HPFP-Bas-mono**) |

| | |
|---|---|
| Behavioural change | Hesitancy and other issues have changed. This has allowed us to have a larger number. Above all, adherence to schedules has also been important. (**IDI01-CSV-Agoè-nyivé**)<br><br>We have observed changes. There have been communities where there are six-year-old children who have never received a dose. (...) The CHWs went to find them. They agreed. When we started, they themselves told us that there were other children too. (...) Some parents are proud to continue so that their children can have the vaccines. (**IDI02-HPFP-Bas-mono**)<br><br>For example, there was one parent who was initially hesitant about vaccination. He had a child who had never been vaccinated since birth (...) But with social marketing, the parent himself took it upon himself to bring the child in and began to follow up. (**IDI03-HFH-Golfe**) |
| **Feasibility** | |
| Simultaneous approach: awareness raising, research and vaccination | (…) the strategy is that when we find the children, we must vaccinate them immediately. (…) when we know that today the ASCs are out in the field going door to door, the vaccination team is on standby. As soon as the children are found, they are vaccinated immediately. (**IDI02-HPFP-Bas-mono**)<br><br>The day before, the targets were identified. On D-day, the CHWs are still there. They go into the houses and bring out the targets they had identified. Also, on D-day, they identify the targets at the same time, the team is there. They vaccinate at the same time. (**IDI04-IFP-Golfe**)<br><br>During vaccination, if we arrive in a region, we vaccinate them there, in the community. We set up a place under the trees or at the notable's or chief's house, and vaccinate them there.<br>For some, we direct them to the USP. But there are certain areas that are a little far from the USP, so whenever we raise awareness, we vaccinate them there. (**IDI06-COGES-Yoto**) |
| Cascade training | First, the stages of CSV training. (...) We tried to train the RFS on all the presentations. And then it was the RFS who went to train the ASCs in our presence. (...) We too had been trained in Atakpamé. (**IDI02-HPFP-Bas-mono**)<br><br>... we were also trained at the beginning. There were ASC training sessions that followed. (...) we also trained them before sending them out into the field. For the community, we had an initial training session for all community members. (...) some members were trained as CSVs. (**IDI03-HFH-Golfe**)<br><br>The training was carried out in a disaggregated, cascading manner. We had to train the regional focal points and DPS. They went on to train the health facilities that carried out the activity. (...) Within the health facilities, they were the ones who trained their vaccinators. (**IDI04-IFP-Golfe**) |
| Home visits | (…) it's a job that's done from house to house. It's door to door, it takes a lot of energy. (IDI01-CSV-Agoè-nyivé)<br><br>... door-to-door visits by CHWs are very important. Because in one day, in a village here, (...) one kilometre from the health centre, the CHW managed to find 25 children. That means it's all good. (**IDI02-HPFP-Bas-mono**)<br><br>For my part, I was informed by a relative who is a community health worker. (...) He came to our house with two of his colleagues, who asked us if our children had been vaccinated. They explained the benefits of vaccination to us. (**IDI07-Beneficiary-Yoto**) |
| Adaptation (necessary adjustments) | ... during the floods, people did it on foot, some did it by canoe. When it rains, some people use canoes to gather children in areas where there is no water. |

| | |
|---|---|
| | As for the insufficient number of CHWs there, we still asked the CHWs who are in the CSV to work as CHWs. We begged the CHWs who are in the monitoring committee to work in the localities as CHWs, even though they are not paid. (**IDI02-HPFP-Bas-mono**)<br><br>For the children who were in class, given that the Ministry of Education was accompanying us, we came to an agreement with them and returned to the hours proposed by the teachers so that we could carry out these activities. This was so that it would not impact their activities as well. (**IDI05-PHD-Vo**)<br><br>For very remote areas, we took motorbike taxis. The USP only has one motorbike, but there was a team that needed to go there, so we took motorbike taxis. (**IDI06-COGES-Yoto**) |
| **Barriers** | |
| Barriers related to sustainability | ... there are some who continue to do their work. There are people who continue to bring children. (...), even though the activity is over, they call us to say they have found this child or that child... They have seen the importance of it. They have seen that if they continue, it can help protect the children in their community. (**IDI02-HPFP-Bas-mono**)<br><br>...given that we had difficulties achieving the current results, we thought that making it permanent would be a good thing. We are in negotiations with the top authorities, the prefect and the mayor. In collaboration with the health authorities, we will see how we can ensure that these teams do not disappear. After this activity, they could continue to work in the field. (**IDI05-PHD-Vo**)<br><br>At present, we have all seen the impact on our activities. So, as of today, what we have done most is to approach local authorities, especially mayors, to see how they can help us to keep our activities going. (...) We are in discussions, and the grievances are on the mayor's table. (**IDI05-PHD-Vo**) |
| Geographical and infrastructural barriers | The main difficulty was that the activity was carried out in rainy weather. And there were difficulties, especially in reaching certain areas. (**IDI05-PHD-Vo**)<br><br>...there are certain parts of our area where sand is mined. This has caused some people to move to these locations. (...) People have moved for work, which has made things a little difficult. But we looked for them. God helped us, and we found them.<br>...only to go to areas that are a little further away, areas far from the USP, the means of transport caused us a bit of trouble. The shortcoming comes from the means of transport that will help us get to where we have encountered difficulties. (**IDI06-COGES-Yoto**) |
| Organisational and resource barriers | But we also noticed that there weren't enough CHWs assigned. (...) There were almost 30 CHWs with community relays. Only 15 were selected. So that bothered us a little. But we tried to manage it that way.<br>And now we can see the number of vaccination team visits. (...) Because in some districts, this has somewhat hindered the number of children vaccinated. Because we have given six visits. And if the CHWs stick to these six visits, there will certainly be children who remain unvaccinated. We need to increase (...) the number of visits by vaccinators. (**IDI02-HPFP-Bas-mono**)<br><br>The difficulty of human resources. Human resources had an impact on implementation and deadlines. This made it impossible to adapt vaccination services to the times desired by the community at all times.<br>In some localities, financial incentives were insufficient, so they had to make do with what they had. (**IDI04-IFP-Golfe**)<br><br>Now, apart from that, it was also school term time. (...) Sometimes it coincided with these children's exams. That's perhaps one difficulty I can mention, given that the target group is at school level. (**IDI05-PHD-Vo**) |
| Community and cultural barriers | ... the main difficulties were caused by the shock of the pandemic. That was the real problem. (...) We had to explain clearly that the vaccines we would be administering were no longer Covid vaccines, but routine vaccines needed to protect children. (**IDI03-HFH-Golfe**) |

| | |
|---|---|
| | It's true that today, we have our beliefs that interfere with this activity in the field.<br>And then there are also social media. (...) And then we also had trouble finding the vaccination records for some children. (**IDI05-PHD-Vo**)<br><br>My children have only had two vaccinations since birth and have not received any other vaccinations to date. When healthcare professionals explain the benefits of vaccination to us, it is convincing. You know, especially the COVID-19 vaccine, it scared people. We were very scared. So I decided not to get vaccinated against COVID-19, and not to vaccinate any of my children against COVID-19. (**IDI07-Beneficiary-Yoto**) |
| **Facilitators** | |
| Awareness raising | This was done at the market. At the market, we tried to set up a podium, a sunshade with the sound system. (...) When mothers come and see that we are vaccinating children at the market (...) this is what made some people returning home from the market bring their health records to the CFWs. This contributed to this. (**IDI02-HPFP-Bas-mono**)<br><br>Through awareness-raising, we give the village dignitary the means to spread the message. After spreading their message, we bring people together and talk to them. (...) We have certain folk groups in our community, as well as tontine groups.<br>This also helps us to find women, as many women are members of tontine groups. If we know when they meet, we visit them in their group. (**IDI06-COGES-Yoto**)<br><br>As my two children were not vaccinated, they offered us the vaccination. (…). They still managed to convince me of the benefits of vaccination and how important it is for our children's lives.<br>When it comes to the message about vaccination, when you go to the hospital, the message gets across. However, when you are at home and people come to visit, they explain things to you in a different way.<br>And even when you are afraid, the fear dissipates. (**IDI07-Beneficiary-Yoto**) |
| Co-creation approach and community engagement | (…) We held meetings with the community. They were the ones who came up with key ideas for mobilising the population around vaccination. (…) It is an advantage to involve people who live in the community (…) they understand that their health is in their own hands. (**IDI03-FHD-Golfe**)<br><br>(…) First, there was a meeting with the authorities, especially local leaders. (…). So, they were the ones who paved the way the most, because when we arrive, it is no longer just the health worker who talks about vaccination, there are also certain leaders who talk and explain to parents and the community the benefits of vaccination. This meant that there was very little hesitancy. (**IDI05-PHD-Vo**) |
| UNICEF-Government Partnership | UNICEF, which spearheaded the social marketing initiative, collaborated with the government. (...) The collaboration consisted of (...) pooling the various services (...) by inviting other sectors that UNICEF considered influential for the success of social marketing, notably the health promotion division and the immunisation division of Ministry of health (...). This was crucial. (**IDI04-IFP-Golfe**) |
| **Recommendations** | |
| | Now we need to try to get this on community radio stations because in our communities, if the information isn't broadcast on the radio, it's as if it's not true. People will tell you that they didn't hear it on the radio. (...).<br>Next time, we also need to go to the IBC (Community-Based Intervention) focal points. In addition to the EPI and Health promotion focal points, they should accompany us in our activities. (**IDI02-HPFP-Bas-mono**)<br><br>(…) We need to go further to ensure the long-term future of the CSV (Vaccination Monitoring Committee). So that it becomes a permanent fixture in the community. (**IDI03-HFH-Golfe**) |
